# The Interplay of asthma fear, avoidant behavior, and Climate Change Emotions: Critical Asthma Syndrome sample

**DOI:** 10.1101/2025.05.26.25328365

**Authors:** Mohammed Musaed Al-Jabri, Shimmaa Mohamed Elsayed, Ahmed Hashem El-Monshed, Mohamed Hussein Ramadan Atta

## Abstract

**Aim:** To assess levels of asthma fear, avoidant behavior, and climate change emotions among patients diagnosed with critical asthma syndrome and explore the relationships between these variables.

**Design:** A multi-center, cross-sectional study adhering to the STROBE guidelines.

**Methods:** A purposive sample of XXX patients with critical asthma syndrome was recruited from intensive care units (ICUs) in general hospitals and educational institutions in Alexandria’s main university hospital and Saudi Arabia. Data collection took place from XXXXX to XXXXX. The study employed three validated instruments: the Fear of Asthma Symptom Scale (FAS), the Asthma Behavior Checklist (ABC), and the Inventory of Climate Emotions (ICE).

**Results:** Participants reported moderate levels of asthma fear and a relatively high degree of avoidant behavior. Climate emotions, particularly anger, and powerlessness, were significantly associated with both asthma fear and avoidant behavior. Regression analysis indicated that asthma fear and climate emotions, especially climate anger and sorrow, were significant predictors of avoidant behavior.

**Conclusion:** Climate change emotions play a crucial role in shaping asthma-related psychological and behavioral responses. Recognizing this relationship can inform psychological interventions to improve asthma management and address emotional responses to climate change among individuals with critical asthma syndrome.

**Implications for the Profession and/or Patient Care:**

- Enhancing asthma management strategies by incorporating psychological interventions targeting fear and avoidance behaviors.
- Addressing climate-related emotions in asthma care may help reduce avoidant behaviors and improve patient outcomes.
- Raising awareness of the emotional impact of climate change on asthma patients can guide future policy and healthcare initiatives.

**Impact:**

- **Problem Addressed:** The study explores how climate change emotions influence asthma-related fear and avoidance behaviors.
- **Main Findings:** Climate emotions, particularly anger and sorrow, significantly predict avoidant behaviors in individuals with critical asthma syndrome.
- **Research Impact:** The findings have implications for psychological interventions, healthcare strategies, and policy development aimed at improving asthma management in the context of climate change.

**Reporting Method:** This study adhered to the STROBE guidelines for cross-sectional studies.

**Patient or Public Contribution:** Patients with acute asthma are the main subjects of our study.

**What Does This Paper Contribute to the Wider Global Clinical Community?:**

- Highlights the need for an integrated approach in asthma management that considers both medical and psychological factors, including climate-related emotions.
- Provides evidence for the role of climate change emotions in shaping health behaviors, informing future interventions in respiratory care.
- Supports the development of patient-centered strategies to reduce asthma-related fear and avoidance through targeted psychological support.

## Introduction

Climate change, driven by both natural processes and human activities, is a significant concern worldwide, profoundly impacting human health and well-being. Natural contributors to climate change include natural greenhouse gas emissions, but human activities such as industrial emissions, deforestation, and the burning of fossil fuels accelerate its progression **(Abbass et al., 2022)**. Climate change is defined by long-term alterations in temperature, precipitation patterns, atmospheric pressure, and humidity levels, disrupting ecosystems and exacerbating health challenges. Its manifestations, including rising global temperatures, receding ice sheets, and unpredictable weather patterns, have far-reaching environmental and societal repercussions **(D’Amato et al., 2023**; **Alahin et al., 2024).**

Among the many health effects of climate change, respiratory health is particularly vulnerable to its impacts. Key pollutants, such as particulate matter (PM2.5 and PM10), carbon monoxide (CO), ozone (O3), nitrogen dioxide (NO2), and sulfur dioxide (SO2), play a significant role in respiratory disorders (Mm, 2024; Schneider et al., 2021). These pollutants are produced by human activities, including industrial emissions, traffic fumes, wildfires, and domestic heating **(Manisalidis et al., 2020).** When inhaled, delicate particulate matter penetrates deep into the respiratory system, triggering or worsening conditions like asthma, bronchitis, chronic obstructive pulmonary disease (COPD), and lung infections. Vulnerable populations, such as individuals with pre-existing respiratory conditions, are disproportionately affected by these pollutants **(Atta et al., 2024; Khan et al., 2023).**

In addition to its physical health impacts, climate change exerts a profound psychological toll. Communities repeatedly exposed to climate-related calamities, such as floods and wildfires, experience displacement, loss of homes, and disruption of social networks, leading to severe emotional distress **(Factors & Service, 2025).** These experiences often result in heightened fear, anxiety, depression, and avoidant behavior, further aggravating the ability to manage health problems effectively. For example, individuals with respiratory illnesses may neglect their treatment or avoid seeking medical care due to stress or psychological burden, thereby worsening their condition **(Atta et al., 2024b; Manisalidis et al., 2020)**. Addressing the dual challenges of climate change and its health consequences requires an integrated approach to mitigate environmental damage while providing adequate support for both physical and mental health **(Intergovernmental Panel on Climate Change (IPCC), 2023)**

## Background

Asthma is a chronic inflammatory disease characterized by time-varying respiratory symptoms, such as wheezing, shortness of breath, and fluctuating expiratory airflow restriction. These symptoms vary in intensity and duration and are often triggered by allergens, irritants, infections, physical activity, or climate change **(Molnár et al., 2021; Panek et al., 2020; Figueiredo et al., 2023)**. In contrast, acute asthma refers to a severe, life-threatening exacerbation of asthma symptoms, also known as Critical Asthma Syndrome (CAS). CAS encompasses conditions like refractory asthma, near-fatal asthma, and status asthmaticus. It is marked by rapidly worsening shortness of breath, paradoxical breathing, progressive respiratory exhaustion, and lightheadedness, requiring immediate medical intervention **(Chen et al., 2022; Chen et al., 2022; Tépach et al., 2019).** The key distinction lies in the chronic nature of asthma with manageable, fluctuating symptoms versus the acute, severe, and treatment-resistant episodes in CAS.**(Figueiredo et al., 2023; Jradi, 2017and Bonnert et al., 2021a)**

Asthma is one of the most prevalent respiratory disorders, affecting 260–334 million people globally, and ranks as one of the top 20 causes of disability **(Mm, 2024**; **Samaha et al., 2015).** The prevalence of this ailment continues to climb, with forecasts predicting an increase of more than 100 million individuals by 2025. **(Global Asthma Network 2018)**. It resulted in around 1,000 fatalities every day. Most of these deaths occur in low– and middle-income nations, and the majority of them are avoidable. Asthma disrupts people’s jobs, schooling, and family lives. It is growing increasingly common in many emerging economies, and the expense of asthma treatment for healthcare systems, communities, and individuals is rising**. (World Health Organization, 2021; Global Initiative for Asthma, 2022; Bousquet et al., 2020; GINA, 2024)** Respiratory irregularities are commonly noted; research has looked at the relationship between climate change and the respiratory system, revealing light on the quantity of CO2 in expired air and irregular breathing. **(Prezzavento, 2024)**

Climate change emotions refer to affective experiences that are closely linked to the climate crisis. However, various factors can influence a person’s emotions at any given time, including their overall life circumstances, temperament, everyday events, social interactions, and the effects of climate change **(Pihkala, 2022).** Climate change elicits many emotional responses, including ‘worry,’ ‘guilt,’ and skepticism. However, other emotions, such as ‘powerlessness,’ ‘anger,’ and ‘confusion,’ have received less attention but are as important in predicting behavioral changes in response to this climate crisis.**(Iniguez-Gallardo et al., 2021)**

The relationship between climate change and asthma is becoming better recognized, mainly through the perspective of climate change emotions, which can worsen asthma symptoms and general health. According to research, climate anxiety has a considerable influence on asthma management and quality of life, suggesting that psychological variables play an important role in controlling respiratory disorders **(Atta et al., 2024b).** Additionally, climate-related diseases, early mortality, various types of malnutrition, and dangers to mental health and well-being are on the rise. Climate risks are an increasing cause of involuntary migration and displacement. These consequences are frequently interconnected, unevenly distributed across and within societies and will continue to be experienced inequitably due to inequalities in exposure and vulnerability. Extreme weather events have caused cascading and compounding health concerns in all inhabited locations, and these risks are anticipated to rise further as the world warms. **(Intergovernmental Panel on Climate Change (IPCC), 2023)**.

Additionally, environmental pollutants and traffic-related pollution are two key factors affecting asthmatics **(Neira & Prüss-Ustün, 2016).** Although research acknowledges the importance of emotions in inspiring climate change action, there are no recommendations for including emotions in Climate Change Education. **(Deisenrieder et al., 2024)** Anxiety disorders are twice as frequent in asthma patients as in the general population, and patients with concomitant anxiety have poorer asthma control, lower quality of life, lower treatment adherence, and higher healthcare utilization**.(Bonnert et al., 2024)**

While the relationship between climate change and respiratory issues, particularly asthma, has been studied in various global contexts, there remains a significant gap in the literature concerning the psychological dimensions of asthma management, specifically Asthma Fear and its impact on avoidant behavior. The current corpus of research has primarily focused on the direct physiological impact of environmental stressors such as increasing temperatures, air pollution, and extreme weather events on asthma exacerbations and treatment. **(Moustafa et al., 2024)** However, limited attention has been paid to the psychological burden that these environmental stressors impose on individuals with asthma, especially in regions heavily affected by climate change. **(Venkatesan, 2023)** Negative emotion sensitivity has been associated with asthma-related distress, and the Negative Emotion Sensitivity Index (NESI) has been utilized in studies on negative emotions and asthma to assess this connection **(Thoren & Petermann, 2000).** However, the NESI is a general measure specifically designed to assess fear of emotional symptoms. Consequently, while the NESI may help identify individuals at risk for emotional disorders within the asthma community, it may not effectively capture distress related explicitly to asthma **(Beckers & Craske, 2017).** Catastrophic thoughts, such as “I do not get enough air,” are common during asthma exacerbations. However, asthma-related distress may also stem from a persistent fear of asthma symptoms, leading to avoidance behaviors. The measure evaluates intense cognitions associated with asthma exacerbations **(Bonnert et al., 2023).**

Asthma and emotional reaction problems have had mixed outcomes. Most trials have evaluated asthma patients with a concomitant panic condition following an established strategy. While there are some encouraging outcomes, additional therapy development and refining are required**. (Bonnert et al., 2021b)** Thus, emotional reactions in asthma may not be tied to a specific anxiety illness, such as panic disorder, but may be influenced by a proclivity to react with worry to respiratory symptoms. If this is the case, the treatment should be expanded to cover those who have subclinical levels of anxiety related to their asthma. The gold standard technique to alleviate anxiety is exposure-based cognitive-behavioral therapy. **(Kew et al., 2016)** Avoidance of risky stimuli has evident adaptive significance for organisms. However, it frequently becomes maladaptive when it generalizes to stimuli that are not threatening, happen over a lengthy period, or incur excessive costs**. (Rogers et al., 2020)** In asthma, avoidant conduct refers to a patient’s inclination to minimize or avoid activities that they believe would trigger or intensify their symptoms. This behavior might include avoiding activity, environmental triggers (such as pollen or smoking), and even social circumstances where asthma symptoms may worsen (for example, crowded locations or areas with poor air quality). This pattern of behavior is frequently perpetuated by earlier experiences of asthma exacerbations when patients learn to identify specific activities or circumstances with pain or worry**.(Rogulj et al., 2024)** The intensity of avoidant conduct determines how it is classed. At a basic level, avoidance might entail taking measures like using inhalers before physical activity or wearing a mask in particular situations. At higher degrees, avoidance can become a crippling disorder when individuals retreat from all potential triggers and drastically limit their day-to-day activities. **(Beckers & Craske, 2017)**

Both fear and avoidance can impair successful asthma control. Fear can cause poor adherence to approved asthma treatment programs because of a lack of faith in the drug or fear about adverse effects. **(Rogulj et al., 2024)**

Furthermore, avoidant conduct may lead to a loss of physical fitness and lung function, which might aggravate asthma symptoms over time. Interestingly, evidence shows that addressing psychological aspects such as anxiety and avoidance in asthma therapy can considerably improve clinical results, including lowering emergency department visits and hospitalizations.**(Atta et al., 2024b; Panagiotou et al., 2020).** In relation to asthma, climate emotions **(Marczak et al., 2024)** have been found to be distinctly associated with climate change perceptions, support for pro-climate policies, socio-demographic factors, loneliness, and alienation, as well as environmental activism and the willingness to prioritize environmental well-being over immediate self-interest. Their study suggests that both positive and negative emotions regarding climate change may be linked to pro-environmental attitudes and behaviors. Additionally, their findings indicate that women and young individuals tend to be more emotionally engaged with climate change. Similarly, **Lawrance et al. (2022)** identified a connection between climate anxiety and pro-environmental decision-making, highlighting that moderate levels of climate anxiety generate an optimal level of arousal, which in turn facilitates an increase in pro-environmental choices.

The gap in the research becomes even more apparent when addressing climate emotional reaction to environmental circumstances, such as asthma fear, and its influence on avoidant behavior**. (Lawrance et al., 2022)** While previous research has identified anxiety as a contributing factor to the worsening of physical health conditions in general, little has been done to investigate how climate-related psychological stress, such as asthma fear and its impact on avoidant behavior, as well as Climate Change Emotions among asthmatic patients affects these conditions**.(Schipper et al., 2024)** Furthermore, current research focuses primarily on demographic and disease-related variables, with little investigation into how the emotional and psychological components of climate change, particularly climate fear, link to asthma outcomes.**(Alahin Arif Salman Yosif Al Bayati, 2024).**

Despite the growing body of research highlighting the relationship between climate change and respiratory issues, particularly asthma, significant gaps remain in understanding the psychological dimensions of asthma management. Most existing studies focus on the direct physiological impacts of environmental stressors, such as air pollution, rising temperatures, and extreme weather events, on asthma exacerbations and treatment outcomes **(Moustafa et al., 2024; Venkatesan, 2023)**. However, limited attention has been paid to the psychological burden imposed by these stressors, particularly in regions heavily affected by climate change. This includes the unique emotional reactions of asthmatic patients, such as asthma fear, and how these emotions influence avoidant behavior and overall asthma control.

Thus, our study aims to address this significant gap by being the first to investigate the relationship between asthma fear, avoidant behavior, and climate change emotions in patients with critical asthma syndrome. This research seeks to explore how these variables interact to influence asthma control, offering a novel perspective on the psychological dimensions of asthma management in the era of climate change.

## 3 | THE STUDY

### 3.1 | Aim of the study

To determine the level of asthma fear, avoidant behavior, and Climate Change Emotions among critical asthma syndrome patients. To explore the interplay of asthma fear, avoidant behavior, and Climate Change Emotions among Critical Asthma Syndrome patients. Three objectives of our study included (1) assessing the level of asthma fear, (2) the level of avoidant behavior, (3) the climate change emotions, and (4) exploring the relationship between asthma fear, avoidant behavior, and climate change emotions among critical asthma syndrome patients.

## 4 | METHOD

### 4.1 | The study design and setting

This study used a multi-center, cross-sectional design following the ‘Improving the reporting of observational studies in epidemiology’ (STROBE) checklist. It included XXX patients diagnosed with critical asthma syndrome. The study sample was recruited from intensive care units of general hospitals and educational institutions in Alexandria’s main university hospital and Saudi Arabia. Data was collected from XXXXX to XXXXXXX.

### 4.2 | Participants

The purposive sample technique was used in the current study. Patients’ inclusive criteria included patients of both sexes, ages from 20 to 80 years. Patients who were admitted to the ICU diagnosed with critical asthma syndrome with the following criteria will be included in this study: (1) inability to speak or only a few words, (2) a reduced peak expiratory flow rate (PEFR) of <25 % of a patient’s personal best, (3) vital signs: tachycardia, and tachypnea, (4) oxygen saturation <90%, (5) altered mental status, may be severed agitation, (6) moderate to severe use of accessory muscles, (7) severe wheezing or decrease breath sound, (8) End-tidal CO2 >35mmHg or Hypercapnia, (9) a failed response to frequent bronchodilator administration and intravenous steroids **(Schivo et al., 2015).**

### 4.3 | Study instruments and measures

#### 4.3.1 | Instruments of data collection

This study employed three instruments in the Arabic language to gather data in addition to demographics and social and clinical data (e.g., gender, age, disease duration, work status, etc.).

##### Socio-demographics and Clinical Data

It includes age, gender, history, marital status, level of education, years of respiratory illness, and family history.

##### Instrument I: Fear of Asthma Symptom Scale (FAS)

It was developed by **Bonnert et al., 2023.** Developed initially with 12 items, the scale was later refined to 11 items. It is a patient-reported outcome measure designed to assess the level of fear related to asthma symptoms. Participants are asked to specify how much they agree or disagree with each statement on a scale ranging from 0 (Do not agree at all) to 5 (Very much agree). A higher score on the FAS indicates a higher level of fear related to asthma symptoms. The internal consistency of the FAS in the sample was excellent, with a Cronbach’s alpha of 0.94 (CI 95%: 0.92, 0.95), suggesting the tool is highly reliable **(Bonnert et al., 2023).**

##### Instrument II: The Asthma Behavior Checklist (ABC)

It was developed by **(Bonnert et al., 2023)**, and the Asthma Behavior Checklist (ABC) was designed to assess avoidant behavior in individuals with asthma. The original checklist consisted of 29 items, which were subsequently refined to 8 key items. The response options were rated on a 7-point scale, ranging from 1 (never) to 7 (always). The ABC-8 demonstrated high internal consistency (α = 0.92, 95% CI: 0.90, 0.94) and good test-retest reliability, with correlation coefficients of scale scores over time (r = 0.88) **(Bonnert et al., 2023).**

##### Instrument III: The Inventory of Climate Emotions (ICE)

Climate Emotions (ICE) was developed in 2023 by **Marczak (2024)**; a self-report measure was designed to assess individuals’ emotional responses to climate change. This scale recognizes the complex nature of climate emotions and the social dimensions of climate change. It consists of 32 items representing eight underlying factors: anger (items 1-4), discontent (items 5-8), enthusiasm (items 9-12), powerlessness (items 13-16), guilt (items 17-20), isolation (items 21-24), anxiety (items 25-28), and sorrow (items 29-32). Each factor is measured using four items, all of which demonstrate an excellent fit. Participants respond on a 5-point Likert scale ranging from “strongly disagree” to “strongly agree,” with higher scores reflecting elevated climate emotions. The final subscales exhibited strong internal consistency, with Cronbach’s alpha values of 0.81 or higher **(Marczak, 2024)**. The Arabic version has shown reliability and consistency across both patient and community samples **(Amin et al., 2024; Khalil et al., 2025).**

### 4.4 | Procedure

The authors have obtained permission to use the intended scales for this study. Before beginning data collection, a pilot study was carried out on 10% of the study subjects; six patients were selected randomly and were not included in the data analysis later. This pilot study aims to evaluate the applicability, feasibility, and objectivity and estimate the time required to answer the questionnaire. No modifications were made according to the results of a pilot study. Patients were interviewed individually after obtaining oral informed consent to participate in the study. The questionnaire was completed within 20–30 minutes according to patient endurance.

### 4.5 | Ethical consideration

The faculty of medicine utilized ethical approval by the Research Ethics Committee (REC), Alexandria University approval (IRB: 0306749) in November 2024. The study’s objective was stated to the participants, assuring them that any data obtained would only be used for research purposes. Furthermore, participants were informed of their freedom to deny participation or withdraw from the study before completing the materials without experiencing any negative consequences. The study requested informed written consent from nurses who volunteered to participate. The consent process was completed online, with each participant electronically confirming their consent before advancing to the survey. The data was anonymized to protect confidentiality and security, guaranteeing that the data was only utilized for research purposes. Access to these data was limited to the research team only.

### 4.6 | Data analysis

Table 1 provides a summary of the sociodemographic and clinical characteristics of the study participants. The mean age of participants was 41.16 years (SD = 11.86). In terms of gender, 63.6% of the participants were female, while 36.4% were male. Most participants were married (57.7%), and the most significant proportion of participants had university-level education (61.0%). Looking at participants’ medical history, more than half (53.1%) reported having respiratory diseases, while 31.1% had cardiovascular diseases. Finally, most participants (64.3%) had been living with respiratory diseases for less than 5 years, with a mean disease duration of 4.40 years (SD = 3.24).

**Table 1.**
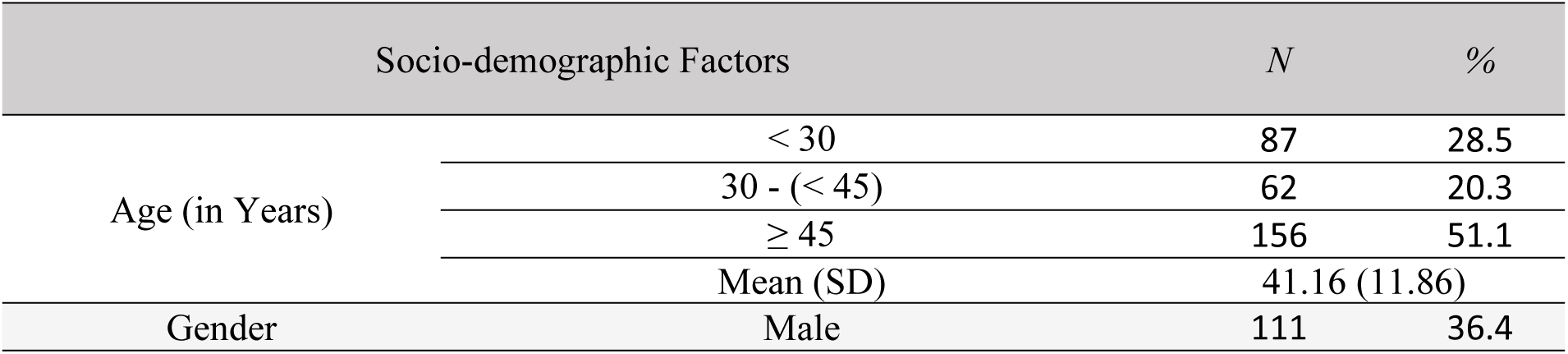
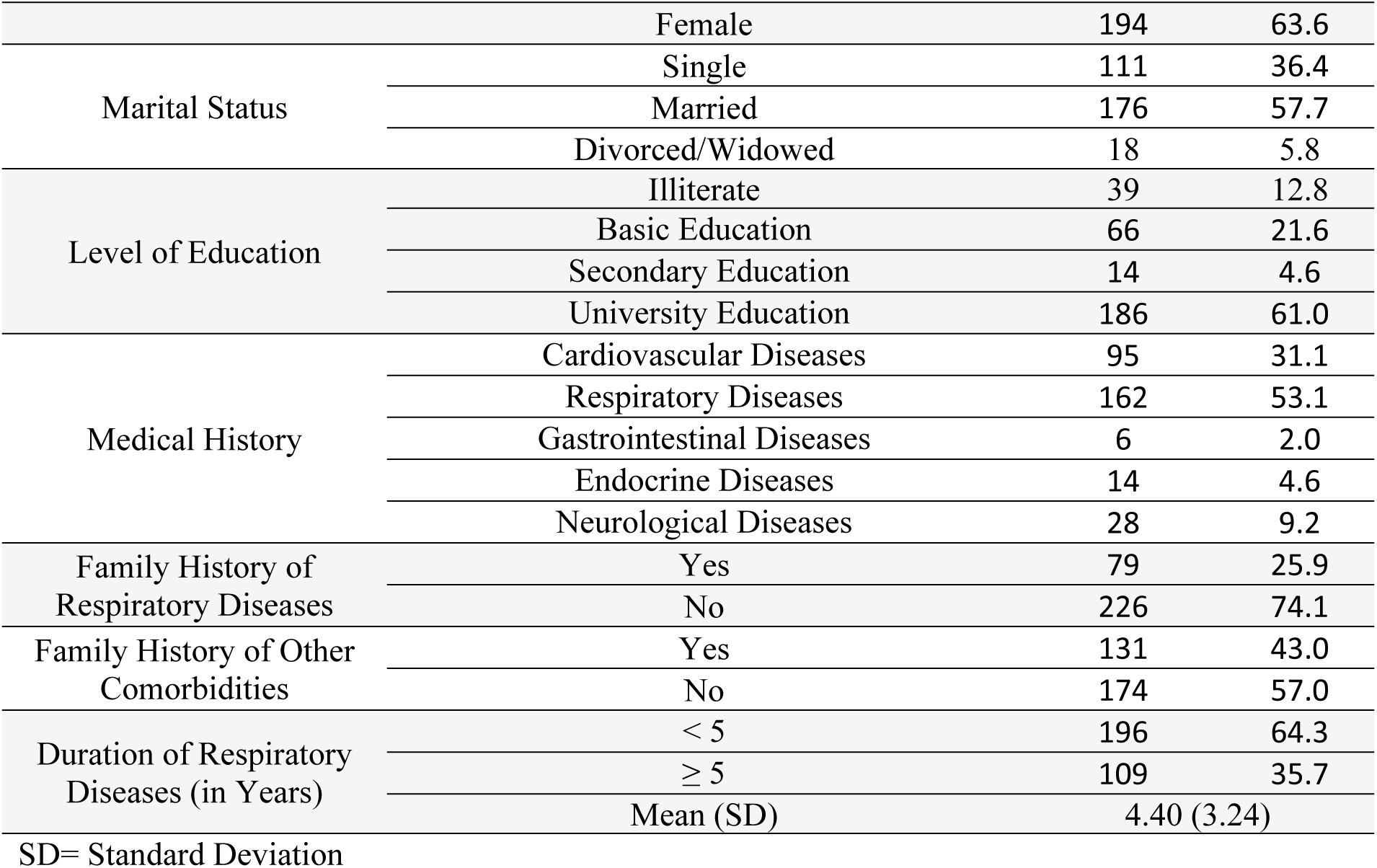
Sociodemographic and Clinical Characteristics of the Study Participants (*N*= 305)

Table 2 summarizes the descriptive statistics for the main study measures among the participants. The Fear of Asthma Symptoms score ranged from a minimum of 10.0 to a maximum of 44.0, with a mean score of 24.58 (SD = 9.35), indicating moderate levels of fear among participants. For Asthma avoidance behavior, scores ranged from 34.0 to 54.0, with a mean of 47.05 (SD = 3.76), suggesting that participants demonstrated a relatively high degree of avoidance behavior related to asthma management. The overall Climate Emotions score had a wide range from 143.0 to 268.0, with a mean of 222.07 (SD = 26.71), reflecting considerable variability in emotional responses related to climate. Breaking down the climate-related emotions, Climate Anger showed a mean of 17.48 (SD = 2.44) out of a maximum possible score of 20, indicating that participants generally experienced high levels of anger. Interestingly, Climate Enthusiasm had the lowest mean score of 5.31 (SD = 2.60), highlighting a typically low level of positive emotional engagement related to climate among participants. In contrast, Climate Powerlessness (M = 16.53, SD = 2.99) and Climate Guilt (M = 16.51, SD = 2.68) both had relatively high mean scores, reflecting strong feelings of helplessness and guilt about climate issues.

**Table 2.** Descriptive Statistics of the Study Measures (*N* = 305)

| Study Measures | Minimum | Maximum | Mean | SD |
| --- | --- | --- | --- | --- |
| **Fear of Asthma Symptom** | **10.0** | **44.0** | **24.58** | **9.35** |
| **Asthma Avoidant Behavior** | **34.0** | **54.0** | **47.05** | **3.76** |
| **Climate Emotions** | **143.0** | **268.0** | **222.07** | **26.71** |
| • Climate Anger | 10.0 | 20.0 | 17.48 | 2.44 |
| • Climate Contempt | 8.0 | 20.0 | 15.21 | 4.11 |
| • Climate Enthusiasm | 4.0 | 20.0 | 5.31 | 2.60 |
| • Climate Powerlessness | 8.0 | 20.0 | 16.53 | 2.99 |
| • Climate Guilt | 7.0 | 20.0 | 16.51 | 2.68 |
| • Climate Isolation | 8.0 | 20.0 | 16.28 | 2.75 |
| • Climate Anxiety | 8.0 | 20.0 | 15.82 | 2.80 |
| • Climate Sorrow | 7.0 | 20.0 | 15.73 | 2.82 |

SD= Standard Deviation

Table 3 displays the results of the correlation analysis between the study measures. Fear of Asthma Symptoms is significantly and positively correlated with Asthma avoidance behavior (r = .575, p < .001). Additionally, Climate Emotions show a strong positive correlation with both Fear of Asthma Symptoms (r = .918, p < .001) and Asthma Avoidant Behavior (r = .680, p < .001). Among the subscales of climate emotions, Climate Enthusiasm is negatively correlated with both Fear of Asthma Symptoms (r = –.300, p < .001) and Asthma Avoidant Behavior (r = –.571, p < .001), suggesting that higher enthusiasm about climate issues is associated with less fear and avoidant behavior. Other climate emotion subscales, including Climate Anger, Climate Contempt, Climate Powerlessness, Climate Guilt, Climate Isolation, Climate Anxiety, and Climate Sorrow, all show significant positive correlations with both Fear of Asthma Symptoms and Asthma Avoidant Behavior, with correlations ranging from moderate to high (p < .001).

**Table 3.** Correlation Analysis between Studied Measures (*N* = 305)

| Climate Emotions |  | Fear of Asthma Symptom | Asthma Avoidant Behavior |
| --- | --- | --- | --- |
| **Fear of Asthma Symptom** | r | 1 | .575** |
|  | P |  | .000 |
| **Climate Emotions (Total)** | r | .918** | .680** |
|  | P | .000 | .000 |
| Climate Anger | r | .625** | .671** |
|  | P | .000 | .000 |
| Climate Contempt | r | .479** | .175** |
|  | P | .000 | .002 |
| Climate Enthusiasm | r | -.300** | -.571** |
|  | P | .000 | .000 |
| Climate Powerlessness | r | .666** | .583** |
|  | P | .000 | .000 |
| Climate Guilt | r | .630** | .621** |
|  | P | .000 | .000 |
| Climate Isolation | r | .894** | .681** |
|  | P | .000 | .000 |
| Climate Anxiety | r | .804** | .695** |
|  | P | .000 | .000 |
| Climate Sorrow | r | .814** | .683** |
|  | P | .000 | .000 |

$r$ = Pearson correlation
\*\*Correlation is significant at the 0.01 level (2-tailed). \*Correlation is significant at the 0.05 level (2-tailed).

Table 4 presents the regression analysis results where Asthma avoidance behavior is the dependent variable, and Fear of Asthma Symptoms, along with subscales of Climate Emotions, are the predictors. The model explains 60.6% of the variance in Asthma avoidance behavior (R² = 0.606, Adjusted R² = 0.594), with an overall significant model (p<0.001). Climate Anger and Climate Sorrow positively predict avoidant behavior (B= 0.296 and B= 2.117, respectively). Conversely, Climate Enthusiasm and Climate Anxiety negatively predict avoidant behavior (B – 0.315 and B –1.767). On the other hand, Fear of Asthma Symptoms, Climate Contempt, Climate Powerlessness, Climate Isolation, and Climate Guilt were not significant predictors.

**Table 4.** Regression Coefficients: Fear of Asthma Symptom and Subscales of Climate Emotions as Predictors of Asthma Avoidant Behavior.

| Predictors | Asthma Avoidant Behavior |  |  |  |  |  |  |
| --- | --- | --- | --- | --- | --- | --- | --- |
|  | Unstandardized Coefficients |  | Standardized Coefficients | *t* | *P* | 95% CI for difference |  |
| | *B* | *SE* | $\beta$ | | | Lower bound | Upper bound |
| **Constant** | 32.490 | 3.242 |  | 10.021 | .000 | 26.109 | 38.870 |
| Fear of Asthma Symptom | .002 | .056 | .004 | .028 | .977 | -.109 | .112 |
| Climate Anger | .296 | .127 | .192 | 2.329 | .021 | .046 | .545 |
| Climate Contempt | -.118 | .061 | -.129 | -1.939 | .053 | -.238 | .002 |
| Climate Enthusiasm | -.315 | .096 | -.218 | -3.280 | .001 | -.504 | -.126 |
| Climate Powerlessness | .200 | .111 | .159 | 1.810 | .071 | -.017 | .418 |
| Climate Guilt | .282 | .151 | .201 | 1.867 | .063 | -.015 | .580 |
| Climate Isolation | -.031 | .323 | -.022 | -.095 | .924 | -.666 | .605 |
| Climate Anxiety | -1.767 | .726 | -1.315 | -2.435 | .015 | -3.195 | -.339 |
| Climate Sorrow | 2.117 | .742 | 1.587 | 2.852 | .005 | .656 | 3.578 |

Effect (B)= Unstandardized Coefficient; $\beta$ = standardized coefficient beta; SE= Standard Error; *t*= Student t test for linear regression
95% CI for difference 95% Confidence Interval for Difference
$R^2 = 0.606$ ( $Adj R^2 = 0.594$ ), $F$ -change = 50.336, $P < 0.001$ .
Dependent variable: Asthma Avoidant Behavior

Table 5 and Figure 1 illustrate the results of the mediation analysis examining the effect of Asthma Avoidant Behavior (AAB) on Fear of Asthma Symptoms (FAS) through Climate Emotions (CE). The Direct Effect model shows a negative direct effect of AAB on FAS after accounting for CE (B = –0.2273, p = 0.0031), indicating that without the mediating role of climate emotions, the relationship changes direction. The Indirect Effect model confirms the mediation effect of CE between AAB and FAS (B = 1.6551, Bootstrapped 95% CI [1.5022, 1.8148]), which is statistically significant. The overall models are highly significant, with strong R² values, highlighting the key role of climate emotions in mediating the relationship between asthma avoidant behavior and fear of asthma symptoms.

**Figure 1.**
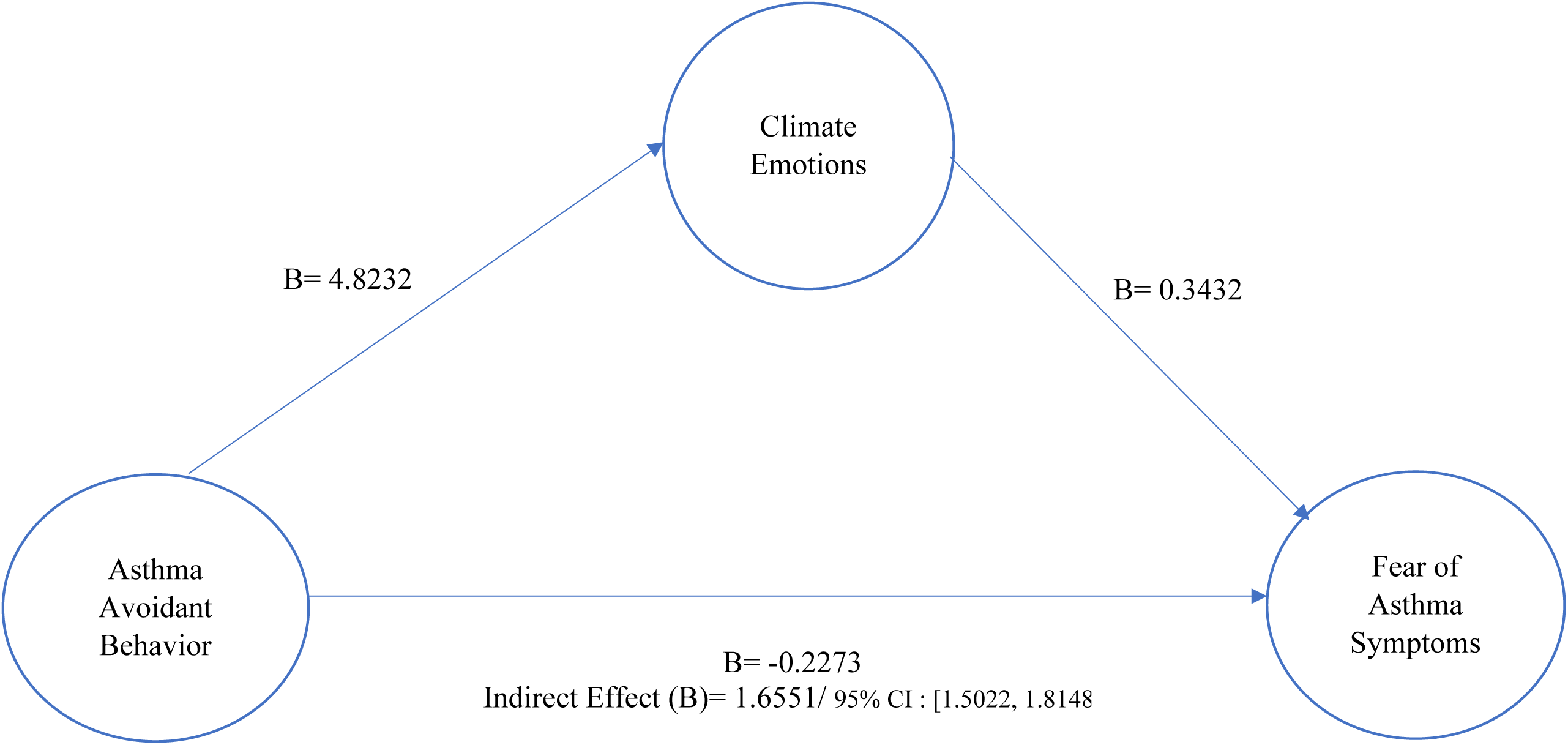
Pathways of Mediation Analysis of the Effect of Asthma Avoidant Behavior on Fear of Asthma Symptoms through Climate Emotions.

**Table 5.** Mediation Analysis of the Effect of Asthma Avoidant Behavior on Fear of Asthma Symptom through Climate Emotions.

| Model/ Path | Effect (B) | SE | t | p | 95% CI for difference |  |
| --- | --- | --- | --- | --- | --- | --- |
|  |  |  |  |  | Lower bound | Upper bound |
| Model 1<br>AAB → CE | 4.8232 | 0.2991 | 16.1240 | 0.0000 | 4.2346 | 5.4119 |
| Model 2<br>CE → FAS | 0.3432 | 0.0107 | 31.9905 | 0.0000 | 0.3221 | 0.3643 |
| Model 3a (Total Effect)<br>AAB → FAS | 1.4278 | 0.1168 | 12.2224 | 0.0000 | 1.1979 | 1.6577 |
| Model 3b (Direct Effect)<br>AAB → FAS | -0.2273 | 0.0761 | -2.9859 | 0.0031 | -0.3772 | -0.0775 |
| Model 3c (Indirect Effect)<br>AAB → CE → FAS | 1.6551 | 0.0806 | - | - | 1.5022 | 1.8148 |
| AAB= Asthma Avoidant Behavior; CE= Climate Emotions; FAS= Fear of Asthma Symptom<br>Effect (B)= Unstandardized Coefficient; SE= Standard Error; *t*= Student t test for linear regression<br>95% CI for difference 95% Confidence Interval for Difference<br>Model 1 Summary for CE (Outcome Variable): $R^2= 0.4618$, $F(1, 303)= 259.9836$, $p= 0.0000$<br>Model 2 Summary for FAS (Outcome Variable): $R^2= 0.8474$, $F(2, 302)= 838.4249$, $p= 0.0000$<br>Model 3 Summary for Total Effect: $R^2= 0.3302$, $F(1, 303)= 149.3873$, $p= 0.0000$<br>Bootstrap Confidence Interval for Indirect Effect: Bootstrapped 95% CI: [1.5022, 1.8148] **(Statistically Significant).** |  |  |  |  |  |  |

## Statistical Analysis

Data analysis was conducted using the Statistical Package for Social Sciences (SPSS), version 29. Categorical variables were summarized as frequencies and percentages, while continuous variables were reported as Mean ± Standard Deviation (SD). Pearson correlation analysis was performed to examine relationships between continuous parametric data. Linear regression analysis was applied to predict the dependent variable based on independent variables. Hypothesis testing included path analysis using the SPSS macro PROCESS (Hayes, 2018). A significance level of 0.05 was maintained for all statistical tests.

## Discussion

Our study has revealed a moderate level of fear associated with asthma symptoms among patients. Similarly, Feldman et al. (2008) examined panic fear, panic disorder (PD), and asthma outcomes among adult asthma patients. They showed that asthma-PD patients experienced greater illness-specific and generalized panic fear compared to asthma-only patients, even though there were no differences in asthma severity or physical symptoms during attacks. Furthermore, Boer et al. (2021) observed a clinically significant increase in anxiety and depression among asthma patients during the pandemic, highlighting the psychological impact of external stressors. Similarly, Deshmukh et al. (2007) reviewed prior research and found an elevated probability of anxiety disorders, particularly panic disorder and panic attacks, in asthma patients compared to the general population. Their review also revealed significant co-morbidity between asthma and anxiety, as measured on dimensional anxiety and panic scales. These findings underline the need for integrated care approaches that address asthma management’s physical and psychological dimensions.

Climate change emotions show a strong positive correlation with the fear of asthma symptoms, likely due to the heightened sense of vulnerability and uncertainty that individuals with asthma experience in the face of climate change. Climate anxiety, a specific form of emotional distress linked to climate-related threats, can exacerbate the fear of asthma symptoms as individuals perceive increased environmental risks, such as pollution, allergens, and extreme weather, which are known triggers for asthma. These emotional responses may be amplified by a lack of perceived control over both the environmental changes and their health conditions, leading to greater fear.

Atta et al. (2024b) provide valuable insights, revealing that climate anxiety was higher among middle-aged individuals, those with longer disease durations, and those with previous hospitalizations. This study also found strong negative correlations between climate anxiety, asthma control, and asthma-related quality of life. These findings suggest that individuals with greater exposure to disease complications or environmental stressors may experience heightened climate anxiety, which undermines their ability to manage asthma and maintain a good quality of life.

Additionally, Janssens et al. (2011) found that fear of symptoms interacts with perceived control over the environment. Individuals with high fear of symptoms experienced more significant unpleasantness during situations where control was reduced, mediated by heightened threat perception. This aligns with the observed positive correlation between climate emotions and fear of asthma symptoms, as those who perceive more significant threats in their environment may experience amplified fear and anxiety about their health. Collectively, these findings underscore the importance of addressing climate emotions and enhancing coping strategies to reduce fear and improve quality of life among asthma patients.

Climate emotions show a strong positive correlation with asthma-avoidant behavior, likely due to the psychological impact of heightened environmental concerns and the perceived threat to health. The findings align with prior research on the effects of climate emotions on asthma-related behaviors and highlight the dual role of emotional responses in influencing avoidant behavior. Negative emotions, such as anger and sorrow, are associated with increased psychological distress and maladaptive coping mechanisms, including avoidance. Individuals with asthma may experience increased emotional distress when they are aware of or directly affected by climate change, such as exposure to extreme weather events or worsening air quality. These emotions, compounded by the fear of symptom exacerbation, may lead to avoidant behaviors as a means of reducing perceived risks. Avoidance strategies, however, may inadvertently limit daily activities and further contribute to emotional and physical health challenges.

Supporting this, Wood et al. (2007) found that emotional climate, or the collective emotional state influenced by environmental and social conditions, was associated with depressive symptoms. These depressive symptoms were linked both directly and indirectly through emotional triggering to the severity of asthma. This highlights how environmental stressors and emotional responses can exacerbate asthma outcomes and encourage maladaptive coping mechanisms like avoidance. Similarly, Lawrence et al. (2022) demonstrated how climate change acts as a risk amplifier by disrupting mental health-supporting conditions, such as stable socioeconomic and environmental environments. The disruptive influence of rising global temperatures, extreme weather events, and climate inaction generates significant psychological distress, particularly among disadvantaged groups, compounding existing stressors.

Our findings reveal that among the subscales of climate emotions, Climate Enthusiasm demonstrates a negative correlation with both Fear of Asthma Symptoms and Asthma Avoidant Behavior, indicating that individuals who feel more enthusiastic about addressing climate issues tend to exhibit less fear of asthma symptoms and less avoidant behavior. This result suggests that positive engagement with climate issues, possibly stemming from a sense of empowerment and active involvement in solutions, mitigates emotional distress and maladaptive responses. Enthusiasm may serve as a buffer against fear by fostering a sense of agency, reducing feelings of helplessness, and promoting adaptive behaviors.

In contrast, other climate emotion subscales such as Climate Anger, Climate Contempt, Climate Powerlessness, Climate Guilt, Climate Isolation, Climate Anxiety, and Climate Sorrow show significant positive correlations with Fear of Asthma Symptoms and Asthma Avoidant Behavior, ranging from moderate to high levels (p < .001). These findings align with previous research indicating that negative climate-related emotions amplify distress and maladaptive responses. For instance, climate anxiety, a prominent emotional reaction to perceived environmental risks, has been linked to heightened fear of asthma symptoms due to the exacerbation of vulnerability and uncertainty. Atta et al. (2024b) demonstrated that climate anxiety was exceptionally high among individuals with longer disease durations, previous hospitalizations, and greater exposure to stressors, contributing to reduced asthma control and diminished quality of life.

The interaction between emotional responses and perceived control further explains this correlation. Janssens et al. (2011) found that individuals with high fear of symptoms experienced more significant distress in situations of low perceived control, mediated by heightened threat perception. This dynamic resonates with our findings, as individuals experiencing emotions like anxiety, guilt, or sorrow may feel overwhelmed by the magnitude of climate-related challenges, leading to avoidance behaviors to reduce perceived threats.

On the other hand, Climate Enthusiasm represents a constructive emotional engagement that counters these patterns. Lawrence et al. (2022) emphasized that while climate change disrupts conditions supporting mental health, strong emotional responses can motivate positive action. Enthusiasm, a forward-looking emotion, likely fosters resilience and a proactive mindset, enabling individuals to perceive challenges as opportunities rather than threats. This sense of agency reduces fear and discourages avoidance, emphasizing the potential of cultivating positive climate emotions to improve both psychological well-being and asthma management outcomes.

Overall, these findings highlight the dual nature of climate emotions, with negative emotions amplifying fear and avoidance and positive emotions like enthusiasm promoting adaptive coping and better health-related quality of life.

Fear of asthma symptoms is significantly and positively correlated with asthma-avoidant behavior, as revealed in our study. This finding suggests that individuals with a heightened fear of experiencing asthma symptoms are more likely to engage in avoidant behaviors to prevent potential asthma triggers or symptoms. One possible reason for this correlation is the psychological impact of fear and anxiety, which may amplify the perception of vulnerability and lead to behaviors aimed at minimizing exposure to perceived risks. Avoidant behaviors, while initially intended to prevent distress or symptoms, can inadvertently limit physical activity, social engagement, and overall quality of life.

Supporting evidence for this result can be found in the study by Bonnert et al. (2021), which examined 30 participants who self-reported anxiety related to asthma. The study found that catastrophizing about asthma was significantly correlated with avoidance behavior, fear of asthma symptoms, and reduced quality of life. This suggests that exaggerated negative thoughts about asthma symptoms may intensify fear and lead to maladaptive coping strategies such as avoidance. Together, these findings emphasize the need for interventions that address both fear of asthma symptoms and avoidant behavior, focusing on cognitive and behavioral strategies to help individuals better manage their asthma without compromising their quality of life.

The mediation analysis reveals the mediating role of climate emotions, transforming the relationship between asthma-avoidant behavior and fear of asthma symptoms, as demonstrated by the significant indirect effect. This aligns with Janssens et al. (2011), who found that individuals with low fear of symptoms experienced less distress in conditions where control was perceived, reinforcing the idea that avoidance might initially alleviate anxiety in some cases.

However, the significant indirect effect of climate emotions indicates that climate emotions amplify the relationship between AAB and FAS. Negative climate emotions, such as anxiety and sorrow, likely heighten the perception of environmental threats, exacerbating fears and reinforcing avoidant behaviors. Atta et al. (2024b) support this perspective, noting that climate anxiety is associated with poor asthma control and reduced quality of life, particularly among individuals with prolonged exposure to disease complications and environmental stressors.

## Strengths & Limitations

This study provides valuable insights into the complex relationship between fear of asthma symptoms, avoidant behavior, and climate emotions, highlighting the psychological aspects of asthma management. The use of a comprehensive set of measures and robust statistical analyses, including correlation, regression, and mediation, offers a deep understanding of these interconnections. Furthermore, the study’s exploration of climate emotions adds a novel dimension to asthma research, emphasizing the importance of emotional responses to both asthma and climate change in influencing behavior.

One key limitation is the lack of randomization, which may reduce the generalizability of the findings to broader populations. Additionally, the study did not assess all potential covariates, such as specific exposure to air pollution or other environmental factors, which could further influence asthma outcomes and emotional responses. Future studies could benefit from a more detailed examination of these factors to better understand the full scope of environmental and psychological influences on asthma management.

## Implication

The findings have important implications for understanding and managing asthma-related behaviors and emotional responses. The strong correlations between fear of asthma symptoms, avoidant behavior, and climate emotions suggest that emotional responses to climate change can exacerbate asthma management challenges. Interventions targeting asthma should consider integrating strategies to address not only fear and avoidant behaviors but also the broader emotional context, including anger, sorrow, and anxiety, which may stem from climate concerns. For example, psychoeducation and cognitive-behavioral interventions could help patients develop healthier coping mechanisms to reduce avoidance and improve self-management. Additionally, the significant mediation effect of climate emotions highlights the need for public health initiatives to address climate-related distress, which could indirectly enhance asthma management outcomes. Policymakers and healthcare professionals should also prioritize emotional support in asthma care, particularly for individuals highly affected by climate-related stressors, to mitigate the psychological burden and improve overall quality of life.

## Conclusion

The study highlights the critical interplay between fear of asthma symptoms, avoidant behavior, and climate emotions, emphasizing the need for integrated interventions addressing both psychological and environmental factors to improve asthma management and patient well-being.

## Data Availability

Data availability Data are available with reasonable consent from the authors.

## References

1. Abbass, K., Qasim, M. Z., Song, H., Murshed, M., Mahmood, H., & Younis, I. (2022). A review of the global climate change impacts, adaptation, and sustainable mitigation measures. Environmental Science and Pollution Research, 29(28), 42539–42559. 10.1007/s11356-022-19718-6

2. Alahin Arif Salman Yosif Al Bayati. (2024). The Impact of Climate Change on Biodiversity and Ecosystem Functioning. Academic International Journal of Pure Science, 2(2), 15–25. 10.59675/p222

3. Atta, M. H. R., El-Sayed, A. A. I., Taleb, F., Elsayed, S. M., Al Shurafi, S. O., Altaheri, A., Abdu almoliky, M., & Asal, M. G. R. (2024). The Climate-Asthma Connection: Examining the Influence of Climate Change Anxiety on Asthma Control and Quality of Life: A Multi-National Study. Journal of Advanced Nursing, 1–18. 10.1111/jan.16513

4. Beckers, T., & Craske, M. G. (2017). Avoidance and decision making in anxiety: An introduction to the special issue. Behavior Research and Therapy, 96, 1–2. 10.1016/j.brat.2017.05.009

5. Bonnert, M., Nash, S., Andersson, E. M., Bergström, S. E., Janson, C., & Almqvist, C. (2024). Internet-delivered cognitive-behavior therapy for anxiety related to asthma: study protocol for a randomized controlled trial. BMJ Open Respiratory Research, 11(1), 1–7. 10.1136/bmjresp-2023-002035

6. Bonnert, M., Roelstraete, B., Bergstrom, S. E., Bjureberg, J., Andersson, E., & Almqvist, C. (2023). The Fear of Asthma Symptoms Scale and the Asthma Behavior Checklist: preliminary validity of two novel patient reported outcome measures. Journal of Asthma, 60(8), 1558–1565. 10.1080/02770903.2022.2160343

7. Bonnert, M., Särnholm, J., Andersson, E., Bergström, S. E., Lalouni, M., Lundholm, C., Serlachius, E., & Almqvist, C. (2021a). Targeting excessive avoidance behavior to reduce anxiety related to asthma: A feasibility study of an exposure-based treatment delivered online. Internet Interventions, 25(June), 0–7. 10.1016/j.invent.2021.100415

8. Bonnert, M., Särnholm, J., Andersson, E., Bergström, S. E., Lalouni, M., Lundholm, C., Serlachius, E., & Almqvist, C. (2021b). Targeting excessive avoidance behavior to reduce anxiety related to asthma: A feasibility study of an exposure-based treatment delivered online. Internet Interventions, 25(February), 0–7. 10.1016/j.invent.2021.100415

9. Chen, C., Sharma, R., Singh, A., Fraser, D. R., & Kilburn, J. (2022). Critical asthma syndrome in trauma patients – A case report and literature review. Trauma Case Reports, 42(November), 100729. 10.1016/j.tcr.2022.100729

10. D’Amato, G., Annesi-Maesano, I., Biagioni, B., Lancia, A., Cecchi, L., D’Ovidio, M. C., & D’Amato, M. (2023). New Developments in Climate Change, Air Pollution, Pollen Allergy, and Interaction with SARS-CoV-2. Atmosphere, 14(5), 1–13. 10.3390/atmos14050848

11. Deisenrieder, V., Oberauer, K., Kubisch, S., Parth, S., Stötter, H., & Keller, L. (2024). Emotions affect learning about climate change–a case study of FFF. International Research in Geographical and Environmental Education, 0(0), 1–20. 10.1080/10382046.2024.2342727

12. Deshmukh, V. M., Toelle, B. G., Usherwood, T., O’Grady, B., & Jenkins, C. R. (2007). Anxiety, panic, and adult asthma: A cognitive-behavioral perspective. Respiratory Medicine, 101(2), 194–202. 10.1016/j.rmed.2006.05.005

13. Factors, C., & Service, W. (2025). January 2025 Los Angeles County Wildfires. January.

14. Feldman, J. M., Siddique, M. I., Thompson, N. S., & Lehrer, P. M. (2008). The Role of Panic-Fear in Comorbid Asthma and Panic Disorder. Journal of Anxiety Disorders, 23(2), 178. 10.1016/j.janxdis.2008.06.005

15. Figueiredo, I. A. D., Ferreira, S. R. D., Fernandes, J. M., Silva, B. A. da, Vasconcelos, L. H. C., & Cavalcante, F. de A. (2023). A review of the pathophysiology and the role of ion channels on bronchial asthma. Frontiers in Pharmacology, 14(September), 1–22. 10.3389/fphar.2023.1236550

16. GINA. (2024). Relatório Principal GINA 2024 – Iniciativa Global para a Asma – GINA (p. 263). https://ginasthma.org/2024-report/

17. Iniguez-Gallardo, V., Lenti Boero, D., & Tzanopoulos, J. (2021). Climate Change and Emotions: Analysis of People’s Emotional States in Southern Ecuador. Frontiers in Psychology, 12(September), 1–13. 10.3389/fpsyg.2021.644240

18. Intergovernmental Panel on Climate Change (IPCC). (2023). Health, Wellbeing and the Changing Structure of Communities. In Climate Change 2022 – Impacts, Adaptation and Vulnerability. 10.1017/9781009325844.009

19. Janssens, T., Verleden, G., De Peuter, S., Petersen, S., & Van den Bergh, O. (2011). The influence of fear of symptoms and perceived control on asthma symptom perception. Journal of Psychosomatic Research, 71(3), 154–159. 10.1016/j.jpsychores.2011.04.005

20. Jradi, H. (2017). on the Circadian Pattern of Melatonin Report. Annals of Thoracic Meedicine, 13(3), 156–162. 10.4103/atm.ATM

21. Kew, K. M., Nashed, M., Dulay, V., & Yorke, J. (2016). Cognitive behavioural therapy (CBT) for adults and adolescents with asthma. Cochrane Database of Systematic Reviews, 2016(9). 10.1002/14651858.CD011818.pub2

22. Khan, J., Ghani, S. A., Ahmad, S. A., & Javed, H. (2023). The Invisible Threat: Urban Pollution’s Silent Assault on Respiratory Well-being. 1–4.

23. Lawrance, E. L., Thompson, R., Newberry Le Vay, J., Page, L., & Jennings, N. (2022). The Impact of Climate Change on Mental Health and Emotional Wellbeing: A Narrative Review of Current Evidence, and its Implications. International Review of Psychiatry, 34(5), 443–498. 10.1080/09540261.2022.2128725

24. Manisalidis, I., Stavropoulou, E., Stavropoulos, A., & Bezirtzoglou, E. (2020). Environmental and Health Impacts of Air Pollution: A Review. Frontiers in Public Health, 8(February), 1–13. 10.3389/fpubh.2020.00014

25. Marczak, M. (2024). Emotional responses to climate change: Exploration, measurement and the role of emotions in planetary health (Issue April).

26. Marczak, M., Wierzba, M., Kossowski, B., Marchewka, A., Morote, R., & Klöckner, C. A. (2024). Emotional responses to climate change in Norway and Ireland: a validation of the Inventory of Climate Emotions (ICE) in two European countries and an inspection of its nomological span. Frontiers in Psychology, 15(February), 1–18. 10.3389/fpsyg.2024.1211272

27. Mm, M. M. (2024). Asthma – a healthcare, environment and green transformation – associated disease Natalia Dąbrowska [ND] Independent Public Complex of Health Care Facilities in Pruszków; aleja Armii Krajowej 2 / 4, Karolina Strus [KS] Independent Public Specialist. 1–15.

28. Molnár, D., Gálffy, G., Horváth, A., Tomisa, G., Katona, G., Hirschberg, A., Mezei, G., & Sultész, M. (2021). Prevalence of asthma and its associating environmental factors among 6–12-year-old schoolchildren in a metropolitan environment—A cross-sectional, questionnaire-based study. International Journal of Environmental Research and Public Health, 18(24). 10.3390/ijerph182413403

29. Moustafa, A. A., Khalaf, M., Elganainy, R. A., Goher, G. H., Khalil, Y. S., Arnous, M. O., Hegazy, A., Mansour, S. R., Arnous, M. O., & Hegazy, A. K. (2024). Assessing the Effects of Climate Change on Terrestrial and Marine Flora and Fauna: A Review. 16(October), 73–100.

30. Neira, M., & Prüss-Ustün, A. (2016). Preventing disease through healthy environments: A global assessment of the environmental burden of disease. Toxicology Letters, 259, S1. 10.1016/j.toxlet.2016.07.028

31. Panagiotou, M., Koulouris, N. G., & Rovina, N. (2020). Physical activity: A missing link in asthma care. Journal of Clinical Medicine, 9(3), 1–19. 10.3390/jcm9030706

32. Panek, M. G., Karbownik, M. S., & Kuna, P. B. (2020). Comparative analysis of clinical, physiological, temperamental and personality characteristics of elderly subjects and young subjects with asthma. PLoS ONE, 15(11 November), 1–16. 10.1371/journal.pone.0241750

33. Pihkala, P. (2022). Toward a Taxonomy of Climate Emotions. Frontiers in Climate, 3(January), 1–22. 10.3389/fclim.2021.738154

34. Prezzavento, G. C. (2024). Catching your breath: unraveling the intricate connection between panic disorder and asthma. Exploration of Asthma & Allergy, 97–110. 10.37349/eaa.2024.00032

35. Rogers, D. G., Protti, T. A., & Smitherman, T. A. (2020). Fear, Avoidance, and Disability in Headache Disorders. Current Pain and Headache Reports, 24(7), 1–8. 10.1007/s11916-020-00865-9

36. Rogulj, M., Vukojević, K., & Lušić Kalcina, L. (2024). A Closer Look at Parental Anxiety in Asthma Outpacing Children’s Concerns: Fear of Physical Activity over the Fear of Drug Side Effects. Children, 11(3). 10.3390/children11030289

37. Samaha, H. M. S., Elsaid, A. R., & Sabri, Y. (2015). Depression, anxiety, distress and somatization in asthmatic patients. Egyptian Journal of Chest Diseases and Tuberculosis, 64(2), 307–311. 10.1016/j.ejcdt.2015.02.010

38. Schipper, C. A., Hielkema, T. W., & Ziemba, A. (2024). Impact of Climate Change on Biodiversity and Implications for Nature-Based Solutions. Climate, 12(11), 1–25. 10.3390/cli12110179

39. Schivo, M., Phan, C., Louie, S., & Harper, R. W. (2015). Critical Asthma Syndrome in the ICU. Clinical Reviews in Allergy & Immunology, 48(1), 31–44. 10.1007/s12016-013-8394-7

40. Schneider, C. R., Zaval, L., & Markowitz, E. M. (2021). Positive emotions and climate change. Current Opinion in Behavioral Sciences, 42, 114–120. 10.1016/j.cobeha.2021.04.009

41. Tépach, C., Acosta, M., & Huerta, J. (2019). Definición de síndromes de asma crítico. Revisión de la literatura. *Alergia,* Asma e Inmunología Pediátricas, 26(3), 84–99. https://www.medigraphic.com/pdfs/alergia/al-2017/al173c.pdf

42. Thoren, C. Ten, & Petermann, F. (2000). Reviewing asthma and anxiety. Respiratory Medicine, 94(5), 409–415. 10.1053/rmed.1999.0757

43. Venkatesan, P. (2023). 2023 GINA report for asthma. In *The Lancet*. Respiratory medicine (Vol. 11, Issue 7, p. 589). 10.1016/S2213-2600(23)00230-8

44. Wood, B., Lim, J., Miller, B. D., & Ballow, M. (2007). Family emotional climate, depression, emotional triggering of asthma, and disease severity in pediatric asthma: Examination of pathways of effect. Journal of Pediatric Psychology, 32(5), 542–551. 10.1093/jpepsy/jsl044

